# THRIVEair: A community-based air monitoring network design in a pollution-burdened Philadelphia neighborhood to advance environmental justice

**DOI:** 10.1101/2025.11.20.25340037

**Authors:** Sheila Tripathy, Lisa Frueh, Karlin Moore, Jaslyn A. Nguyen, Sonya Sanders, Sanija L. Aikens, Carol White, Carol A. Foy, Debbie M. Robinson, Mark Clincy, Christal Heath, James Mullison, Peter Winslow, Craig Johnson, Grace Tiegs, KC Wahl, Leah Johnston, Nancy Johnston, Jane E. Clougherty

## Abstract

**Background:** In 2019, an explosion at the Philadelphia Energy Solutions Refinery, one of the largest urban oil refineries in the U.S., led to its shuttering and transition into redevelopment. Local fenceline communities, previously impacted by refinery operations, expressed concern about air toxics, particularly benzene, released during the decommissioning process. To monitor volatile organic compounds (VOCs), including benzene, in fenceline communities, we created THRIVEair, a partnership between environmental justice organization Philly Thrive and Drexel University scientists. Key goals of the project included community-responsive air monitoring, data democratization, and timely report-back of results.

**Methods:** Through an action-reflection-action approach, we co-designed a one-year VOC monitoring campaign from June 2023-June 2024, and data dissemination products, including fact sheets, Teach-Ins, and a public website. We monitored 37 VOCs using one-week integrated samples collected using passive thermal desorption tubes. Nine stationary sites were monitored weekly, and 11 additional sites were monitored on a rotating basis for two one-week sessions in summer and winter.

**Results:** On average, we found that benzene concentrations were relatively low over the one-year monitoring period (mean for stationary sites: 1.32 µg/m^3^, range = 0.30 – 9.04 µg/m^3^), though spatial and temporal variability were evident.

**Conclusion:** Through the air monitoring campaign design and implementation process, THRIVEair supported Philly Thrive’s goal of establishing an air monitoring network in neighborhoods impacted by the former refinery. By providing publicly available air quality data, THRIVEair results can be leveraged in Philly Thrive’s advocacy efforts.

## Introduction

On June 21, 2019, a fire and explosion at the Philadelphia Energy Solutions (PES) refinery released approximately 676,000 pounds of hydrocarbons,^1^ along with 3,271 pounds of hydrogen fluoride into the air and catapulted bus-sized debris across the Schuylkill River.^2^ The refinery was shut down and purchased in 2020 by Hilco Redevelopment Partners (HRP), and is currently undergoing decommissioning and redevelopment into a logistics and warehousing center. It is the largest parcel of private property in Philadelphia and represents 2% of the city’s land.^3^

Despite being shut down, the former refinery released the second-highest benzene emissions among all US refineries in 2020 and 2021, adding to the cumulative burden of air pollution that communities in South/Southwest Philadelphia already experience, that include an international airport, shipping port, multiple highways, and other industrial sources. In 2020 approximately 117,000 people lived within one mile of the former PES refinery, and most fenceline residents are low-income people of color, highlighting environmental justice implications.^4^

Industries such as oil refining, along with common urban sources of air pollution such as traffic, are significant sources of volatile organic compounds (VOCs). VOCs are a large class of chemicals with varying toxicity levels, depending on the compound. One VOC, benzene, is a known carcinogen (e.g., leukemia), that impacts multiple organs including the immune system, reproductive system, and bone marrow/blood cell development.^5^ Benzene often co-occurs with toluene, ethylbenzene, and xylene, collectively known as BTEX.^6^ In the years preceding the June 2019 explosion, PES was responsible for ∼90% of benzene emissions from point sources in Philadelphia, and BTEX concentrations decreased with distance from the site.^7^ Studies indicate that residents living near petroleum refineries have a higher risk of cancer.^8^

When PES was an active refinery, benzene was monitored at 36 sites along the site’s fenceline, as required by the Environmental Protection Agency (EPA).^9^ Starting from the 2019 explosion and through decommissioning, fenceline concentrations decreased overall. However, annual rolling averages remained out of compliance with the EPA action level (9 µg/m^3^ moving annual average) (**Figure 1**, Supplemental **Figure S1**). Exceedances of the action level are not considered violations, but refineries are required to conduct a root cause analysis of the exceedance and take corrective actions to reduce emissions.^10^ Importantly, benzene was not monitored in neighboring communities during this time, except for one government regulatory EPA Air Quality System (AQS) monitor, which measures benzene for one 24-hour period every six days. Notably, benzene was not monitored on the day of the 2019 explosion. Fenceline monitoring stopped in 2022.^11^

**Figure 1.**
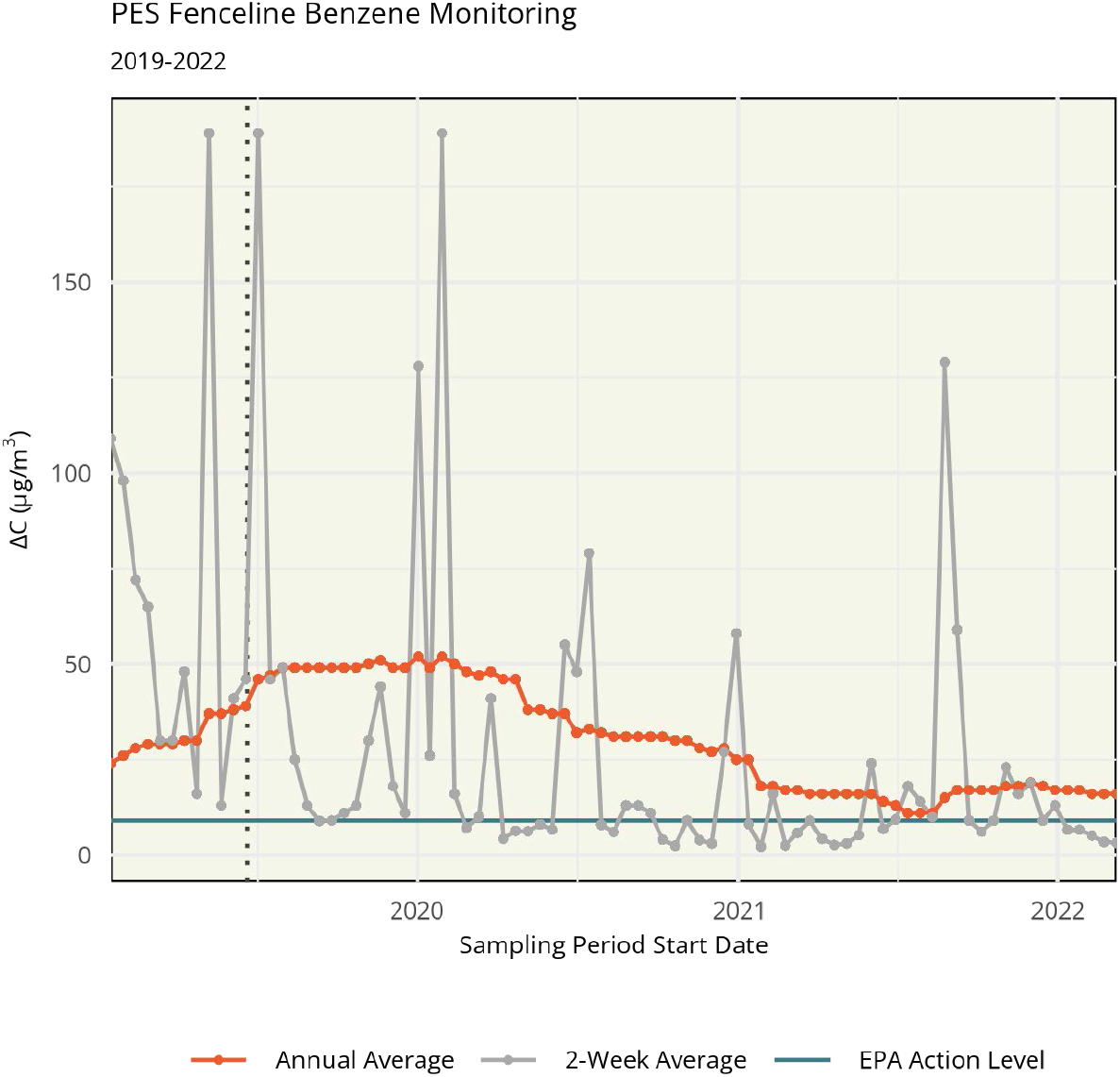
Philadelphia Energy Solutions (PES) refinery fenceline benzene monitoring concentrations from 2019 to 2022. ΔC represents the difference in benzene concentration between the highest- and lowest-measured monitors in a sample period (correcting for urban background levels). The date of the PES refinery explosion is shown in a vertical dotted line. The EPA action level of 9 µg/m^3^ (shown in turquoise) applies to the rolling annual average ΔC (shown in orange).

Since 2015, local environmental justice organization Philly Thrive has been campaigning to shut down the refinery and center environmental justice principles in its redevelopment. By forming a partnership between Drexel scientists and Philly Thrive, THRIVEair aimed to achieve one of Philly Thrive’s goals of establishing a benzene monitoring network in refinery fenceline communities.

### Philly Thrive

Philly Thrive is an environmental justice grassroots organization that has been organizing for the closure of PES and the *Right to Breathe* for all Philadelphians since 2015. Philly Thrive is a multiracial, intergenerational organization that includes members from a wide range of diverse racial and ethnic backgrounds, social classes, income levels, and educational backgrounds. Thrive is a citywide organization, though communities most impacted by refinery operations in South & Southwest Philadelphia represent its primary constituency. *Thrive* is committed to supporting these residents in creating real solutions to environmental change and the ongoing climate crisis through leadership.

After the refinery exploded and was shut down, Thrive established the *Right to Thrive* campaign, which advocates for residents throughout the redevelopment of the site, including cleanup, job creation, and neighborhood investment.^12^ One focus of the *Right to Thrive* campaign was to develop and negotiate a community benefits agreement (CBA) with HRP, including terms related to air monitoring. A community benefits agreement is a legally binding agreement between a developer and one or more community organization/s.

Philly Thrive is organized into smaller groups of “circles” that work on specific goals. The Clean Up Circle was formed to combine members’ lived experience of being exposed to pollution from the site for decades with technical knowledge and research, which was then used to inform advocacy in meeting with the city – and later, HRP - as well as share knowledge about the state of the clean-up with residents. As part of their work, the Circle developed a list of contaminants of concern, including benzene.

Independent air monitoring in South Philly has been one of Thrive’s long time goals. More recent events following the explosion that lead to the THRIVEair project (see timeline in **Figure 2, Figure S2**) included a collaboration with Drexel scientists through a Teach-In event, Thrive’s nonviolent direct action in response to the city’s approval of a permit for the continued use of a tank farm, and an end to fenceline benzene monitoring on the site. Thus, THRIVEair was directly responsive to community calls for continued monitoring of benzene.

**Figure 2.**
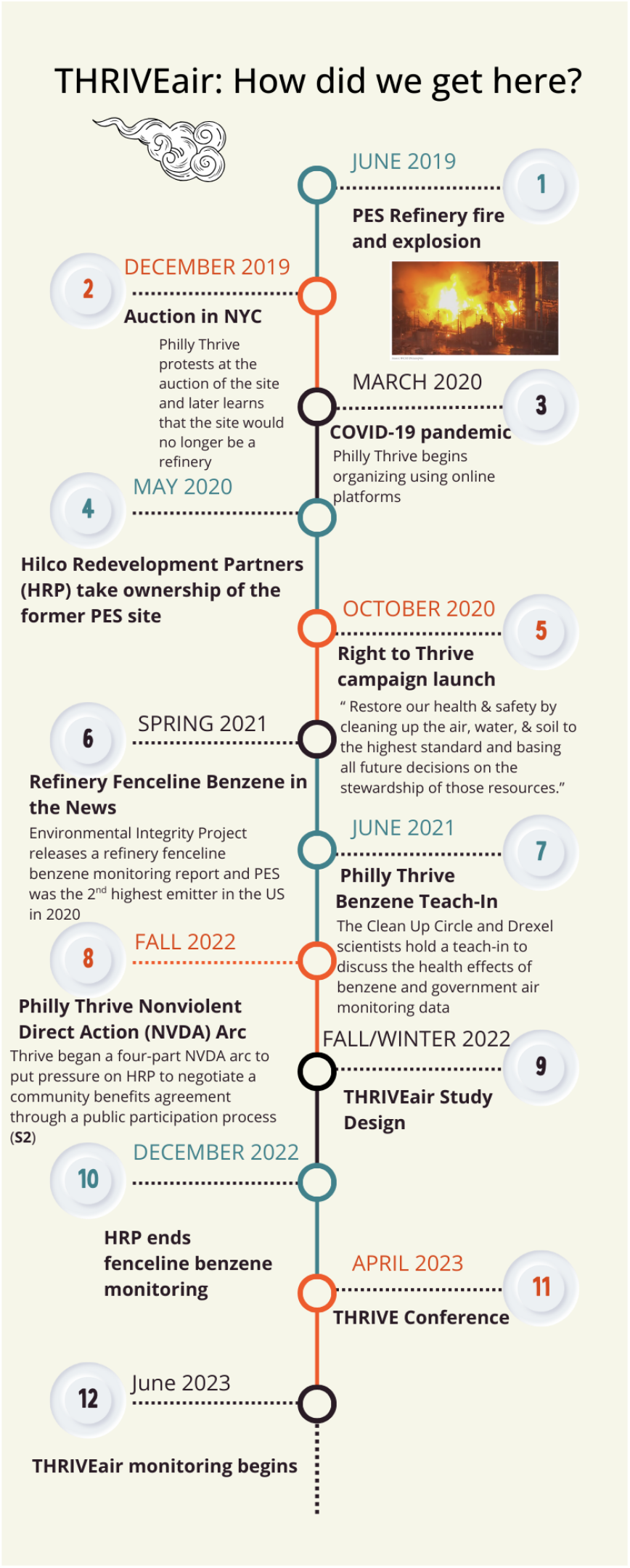
Timeline of Philly Thrive’s organizing activities related to benzene emissions from the former PES refinery and the formation of THRIVEair (For image with photographs please contact first author).

In 2022, Clean Up Circle members and Drexel University public health scientists began planning THRIVEair, a project aimed at establishing an air monitoring network near the former refinery site. Our two main goals were to: 1) Co-design an air monitoring network to monitor VOCs and other air pollutants of concern in neighborhoods adjacent to the former refinery site to help accomplish the “Right to Thrive” campaign goal related to the cleanup of the site and 2) Disseminate results in an ongoing process using methods informed by the Clean Up Circle’s experience on the receiving end of government and corporate public participation processes.

## Methods

### THRIVEair Circle

Our air monitoring campaign design team (the “THRIVEair Circle”) consisted of eight Thrivers, five of whom identified as fenceline members (residents in neighborhoods adjacent to PES), and three public health scientists (one scientist who is also a longtime non-fenceline Thriver, one doctoral student, and one master’s student). Through the report-back co-design process, we engaged three additional fenceline Thrivers, and during the air monitoring campaign, two additional youth Thrivers assisted with air monitoring.

### Air Monitoring Campaign Co-Design

THRIVEair sought to work within Philly Thrive’s established organizing structures as much as possible. Therefore, we engaged the Cleanup Circle in the design process for the air monitoring campaign. In THRIVEair Circle meetings, we based our design process on the praxis approach of “action-reflection-action”, where we held iterative meetings with time for reflection and critical thinking between meetings.^13^ All THRIVEair study design meetings were held in Fall 2022 online on Zoom, due to COVID-19 safety concerns, and participants were compensated for their time. Three initial meetings were held to develop study goals, define the study domain, and select sites. We held an additional meeting open to all Thrivers once we reached a consensus on the study design among the THRIVEair team. We used a combination of methods to organize and develop our project, including examining maps and potential site photos together as a whole group. We also used story sharing rooms (breakout rooms) to work in smaller groups (e.g., creating fenceline-specific story sharing rooms to hear about locations of concern within Grays Ferry and Wilson Park). ^14^^15^

#### Developing Study Goals

In meeting 1, Drexel scientists presented information on air pollutants of concern, including pollutant trends since 2018, using existing government data (e.g., **Figure 1**, Supplemental **Figure S1**). The goal of this was to democratize data that had been challenging to uncover or interpret from government websites. Thrivers highlighted benzene as a primary pollutant of concern and identified the former refinery site and traffic as primary sources of concern.

#### Defining the Study Domain

In meeting 2, Drexel scientists presented several options for study domains for community air monitoring including census tract boundaries that at least partially overlapped with Philadelphia’s city-provided neighborhood maps for Grays Ferry and Wilson Park. After presenting several maps, fenceline members discussed which map most accurately encompassed their neighborhoods without including recently gentrified areas or areas with few residents (e.g., a local shopping center) (Supplemental **Figure S3**).

#### Selecting Community Monitoring Sites

To determine sites of community concern for placing community monitors, we discussed potential monitoring sites as a large group and used story-sharing breakout rooms. Thrivers indicated that parks and schools were sites of concern and pointed out additional locations where community members regularly report chemical odors. Then, Drexel scientists created a web app with potential community sites to guide additional conversations to refine site selection. After compiling potential sites, Drexel scientists stratified the community study domain by traffic density (low vs. high traffic, defined by the 30th/70th percentiles) and distance to the refinery fenceline (near / mid / far), ensuring there were at least two community sites per source class (Supplemental **Figure S4**). After the THRIVEair Circle reached consensus on a monitoring plan, we held a meeting open to all members of Philly Thrive to get more input and finalize our design.

#### Selecting Additional Monitoring Sites

In addition to sites within the Grays Ferry and Wilson Park community domain, we selected one monitoring site upwind from the refinery and five sites in a transect through the middle of and continuing downwind from the refinery to isolate the refinery source effect. We also included one site co-located with a government monitor to assess validity. Three additional sites of community concern were included following discussions within the THRIVEair Circle. However, they were outside of the community study domain: one near a tank farm on the southwest corner of the refinery site, one on a high-traffic street, and one at the former site of Passyunk homes, a location of high community concern.

#### Final Site Selection

Since air monitors were mounted on poles owned by a local electricity company, the final sites were selected as close as possible to the sites determined in the design process, with adjustments made for access feasibility and permit-related issues. The resulting air monitoring locations are shown in **Figure 3**.

**Figure 3.**
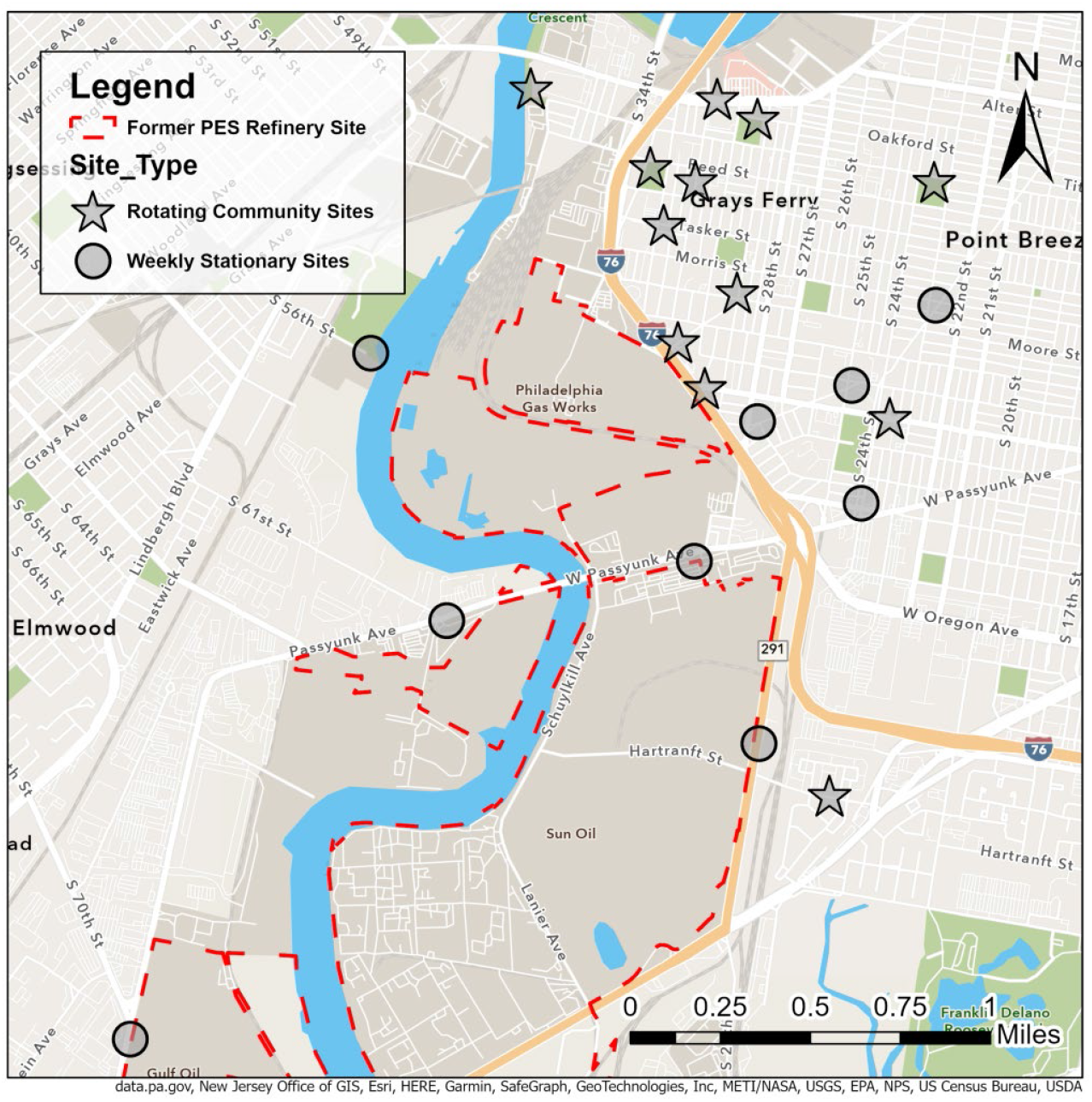
THRIVEair monitoring locations.

#### Outreach

In study design meetings, Thrivers expressed that community members may be suspicious of air monitors on telephone poles in their neighborhood. Therefore, we designed an information tag to hang underneath each monitor (Supplemental **Figure S5**), featuring the THRIVEair project lead’s contact information, a QR code, and a URL for the study website. Throughout the project, Drexel scientists had phone conversations with concerned neighbors and were able to answer questions directly.

### Air Monitoring Methods

#### Air Pollutants

We monitored 37 VOCs, PM_2.5_, and metal constituents at select sites. Here, we present an overview of results related to benzene and benzene derivatives – complete results are reported elsewhere (e.g., the THRIVEair data dashboard and forthcoming publications).

#### Data Collection

VOC data collection ran from June 2023 to June 2024. We mounted Markes passive thermal desorption tubes (TDT) in stainless steel shelters 8-10 feet from the ground on telephone poles, collecting one-week samples with simultaneous deployment and retrieval on Wednesdays. We collected lab and field blanks for each sampling period (2 weeks) and co-located two samples at one site during each sampling session (1 week). We hired a youth Philly Thrive member as a field technician to work with Drexel scientists to deploy and retrieve monitors each week. Every two weeks, the TDTs were shipped to Dr. Johnston’s lab at Lewis and Clark State College for analysis. To prevent sample degradation, TDTs were stored in the refrigerator until they were shipped overnight delivery. We used an electronic field log to track samples. We chose this method of monitoring VOCs because the method is comparable to data collected from the EPA fenceline benzene monitoring program.^16^ Quality control was ensured through the use of field and lab blanks, as well as one duplicate site, for each monitoring week.

#### Laboratory Analysis

Samples were analyzed using thermal desorption gas chromatography with paired mass spectrometry (TD-GC-MS). Quality control was ensured through the use of method blanks and a 5-point calibration for all VOC compounds, including routine standard checks using both gas and liquid standards. Complete laboratory methods are published elsewhere.^17^^18^ Values below the detection limit but above zero were retained and labeled as LOD (limit of detection). Values of zero were replaced with the limit of detection divided by two and labeled non-detect (ND). Values above the upper level of detection were replaced with the upper limit and labeled as the limit of detection (ULOD). Average values of lab and field blanks were subtracted from the raw data. We converted the raw amount (in volume) to parts per billion (ppb) or micrograms per cubic meter (μg/m^3^) using public diffusive uptake rates.

### Report-Back Co-Design

The process of designing materials and methods for data report-backs and dissemination began in Fall 2023. THRIVEair Circle members met over Zoom to discuss data dissemination plans. We reached a consensus to report-back results through a combination of fact sheets, Teach-Ins, and an interactive data dashboard. Thrivers emphasized fact sheets as opportunities to communicate monthly study updates, Teach-Ins as opportunities to connect with the broader community and leverage a dissemination method already employed by Philly Thrive, and a data repository as an opportunity for data democratization.

#### Fact Sheet Co-design

Following an initial meeting on report-back design, Drexel scientists created a fact sheet prototype. Subsequently, two “working group” meetings were held—open to all Thrivers who were compensated for their time. A Thriver was trained as a co-facilitator in these meetings. At the first working group meeting, participants learned about the THRIVEair project and discussed goals for disseminating results. The fact sheet prototype was presented and discussed. Following the first meeting, we developed a revised prototype incorporating the information gathered during the meeting. Working group insights emphasized the use of color, large font for key results, concise “bottom-line” statements, explicit language on health effects and how to interpret concentration units, and a preference for maps over other plot types. A second working group was convened to review and approve these changes, as well as to discuss the website and data repository. The final fact sheet template (Supplemental **Figure S6**) was used to generate monthly updated fact sheets for benzene and BTEX (benzene, toluene, ethylbenzene, and xylenes).

## Results

### Air Monitoring Results: Benzene

The average benzene concentration across stationary sites (which were monitored every week) from June 2023 to June 2024, was 1.32 µg/m^3^ with a range of 0.30– 9.04 µg/m^3^, with substantial variation between sites (**Figure 4**). The site with the highest concentrations was 28th and Passyunk, with an average concentration of 2.05 µg/m^3^ and a range of 0.40–9.00 µg/m^3^. Bartram’s Garden, a site located upwind of the refinery site and Grays Ferry community, had the lowest benzene concentrations (mean 0.92 µg/m^3^). In February 2024, benzene concentrations were elevated across all sites during the week of February 7 to 14, 2024, with higher concentrations at sites closer to the former refinery site. A distance decay was observed, with decreasing concentrations across increasing distance from the site, suggesting an on-site emissions source.

**Figure 4.**
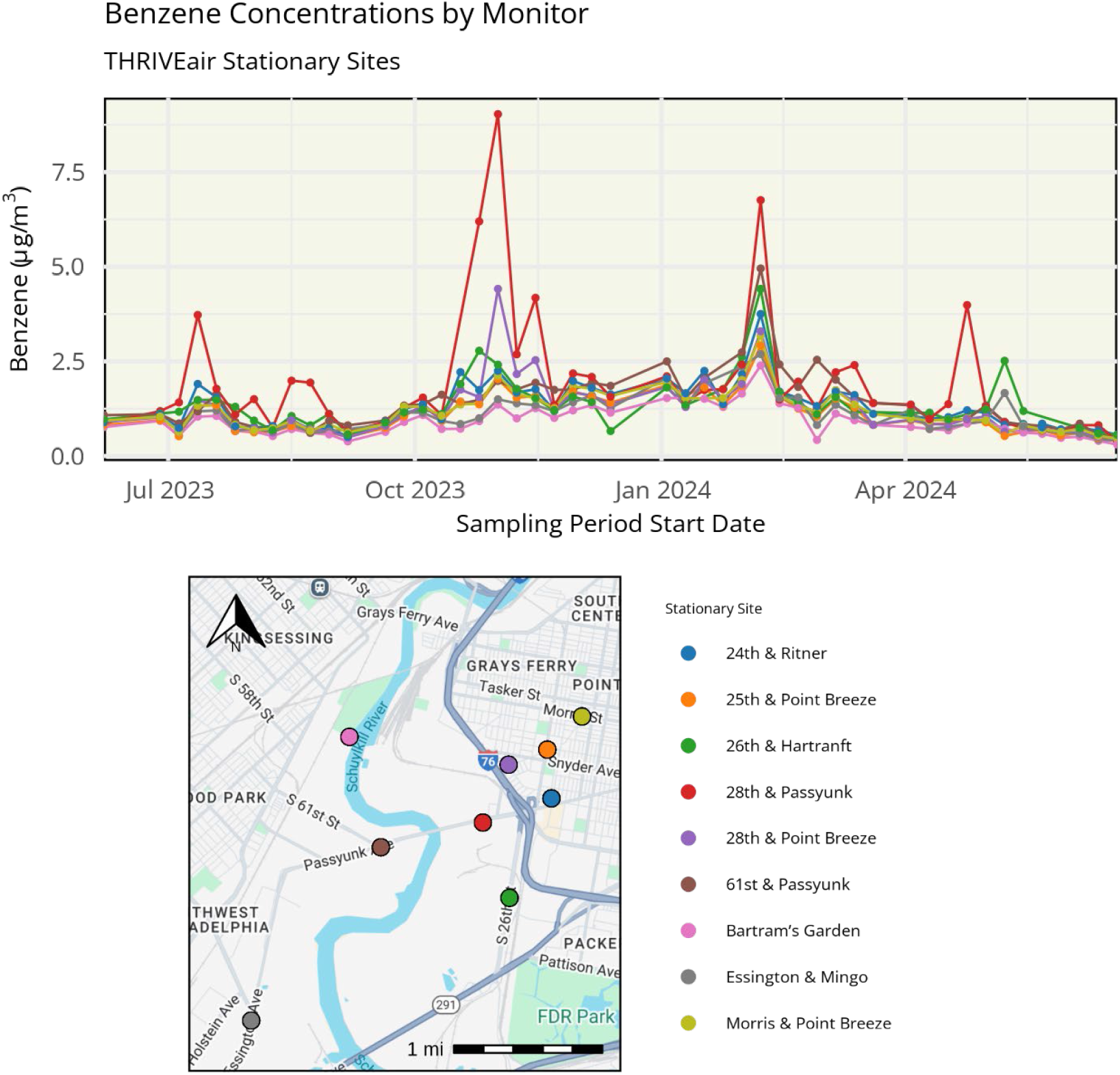
THRIVEair benzene concentrations from June 2023 to June 2024 at stationary sites; bottom) Site locations with matching color codes.

Results from community site monitoring in the summer and winter seasons are shown in **Figure 5**. Benzene concentrations were generally lower at community sites farther away from the refinery compared to stationary sites near the refinery. Full air monitoring results, including temporal and spatial variability, as well as source apportionment, will be published elsewhere (manuscript in preparation), and a link to the manuscript will be provided at thriveairphilly.com.

**Figure 5.**
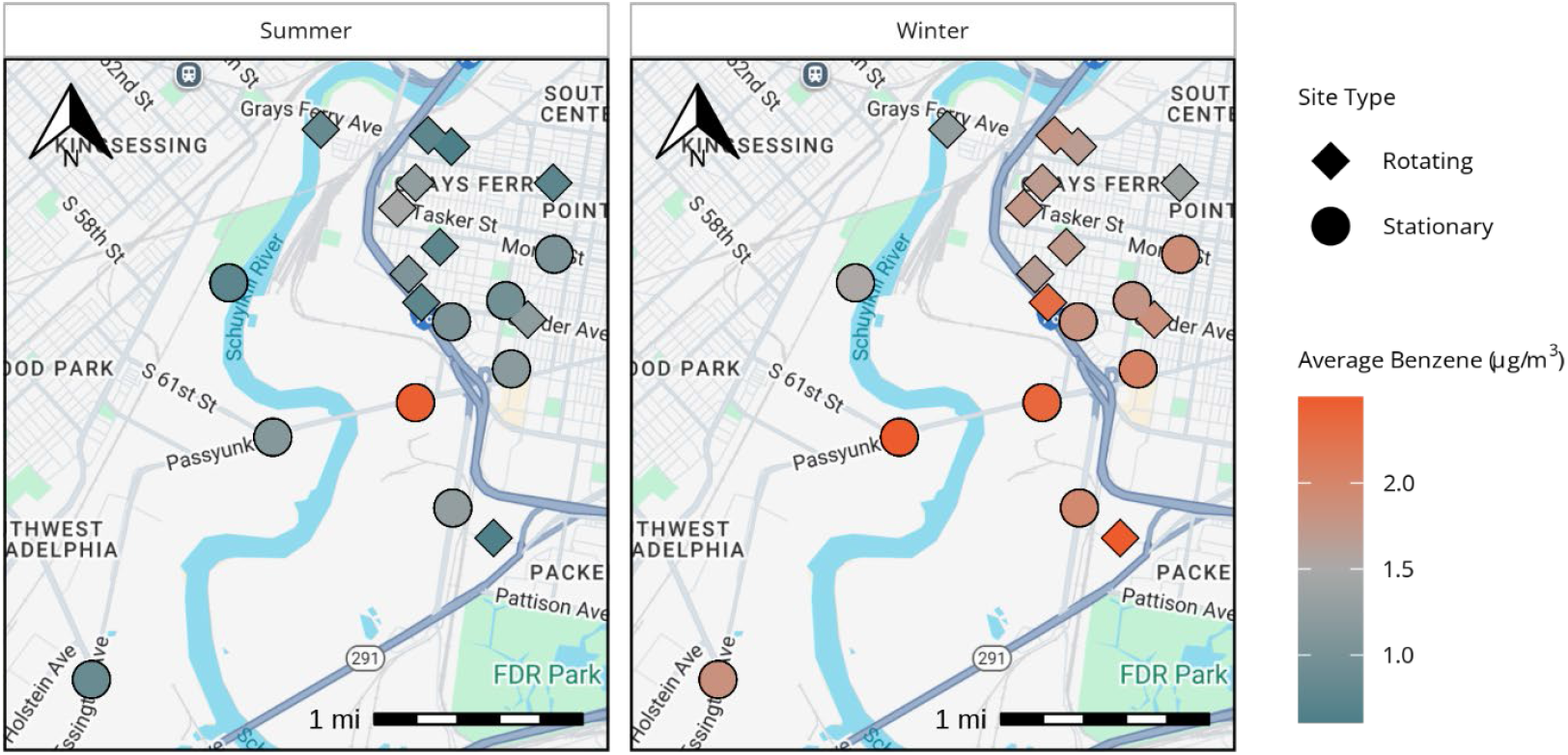
Map of average benzene concentrations for all sites in summer (left) and winter (right) sessions.

### Data Dissemination and Report-Back

Monthly fact sheets describing recent results for benzene and BTEX were printed and placed at Philly Thrive headquarters and posted on the study website, thriveairphilly.com. Additionally, an overall study design fact sheet was distributed (Supplemental **Figure S7**). In June 2024, we presented results, including updated fact sheets, to community members at a Philly Thrive meeting. Our public website, thriveairphilly.com, features an overview of our project, monthly and annual fact sheets, and a data repository. In an interactive R Shiny app, users can visualize results using interactive maps, bar charts, and time series plots. Users can also filter data by pollutant and time range, and data are downloadable in multiple formats (Supplemental **Figure S8**).

#### Outreach

A key aspect of this project was to work within Philly Thrive’s established organizing structures whenever possible during our outreach process. Our first outreach event took place at Philly Thrive’s conference, “Healing the Land and Repairing Harm: A Summit on Philly’s Refinery Redevelopment,” in April 2023. Drexel students set up a table during lunch to showcase the air monitoring equipment to community members and engage in conversations about the project and its study design. We held a “Teach-In” at Philly Thrive’s October 2023 Circle Fair to discuss our study and initial results with community members. We also posted information specific to THRIVEair on Philly Thrive’s membership Facebook group, as well as on Philly Thrive’s public Facebook and Instagram pages and stories.

Towards the end of our study in June 2024, we presented results, including updated fact sheets, to community members at a Philly Thrive meeting. We collaborated with Philly Thrive’s Mutual Aid Circle to include our study factsheet in 150 bags that were distributed to community members at a Philly Thrive meeting, which also included children’s books, environmentally friendly cleaning supplies, and COVID-19 tests.

### THRIVEair Results as a Tool for Advocacy

THRIVEair monitoring results helped inform the direction of the *Right to Thrive* campaign goals. For example, the relatively low concentrations of benzene found among community sites informed a shift in campaign focus to underground plumes of benzene and occupational health issues related to construction workers on site.^19^

While we measured low benzene concentrations on average (mean = 1.32 µg/m^3^), there were several weeks with elevated levels of benzene, with the highest one-week average concentration found at 28th and Passyunk, at 9.04 µg/m^3^. Concerned with these peaks, two members of THRIVEair met with HRP representatives to ask about what these spikes might be related to. Although no information on the source of the spike concentrations was reported, HRP agreed to start monitoring benzene at a site on their property near the Schuylkill River tank farm. The tank farm remains a location of community concern due to the potential use of fossil fuels on the site. HRP began monitoring benzene using a similar sampling protocol to THRIVEair, which helps aid in data comparison with our results. They have continued to monitor benzene at this site and provide us with weekly benzene concentrations, which are also shown in our data dashboard at thriveairphilly.com.

## Discussion

THRIVEair helped accomplish a long-standing Thrive goal of establishing an independent air monitoring network in the neighborhoods surrounding the former PES refinery and providing publicly available air quality data that Thrivers trust and can use to advocate for cleaner air. We conducted a full year of weekly air monitoring and designed a report-back method including a website with a data repository, disseminated factsheets (monthly and for the entire study period), and presented results at Philly Thrive events.

Importantly, THRIVEair results were directly applicable to the advocacy efforts of Philly Thrive. Our findings of relatively low concentrations of ambient benzene in communities neighboring the former refinery site during early stages of redevelopment shifted the *Right to Thrive* campaign environmental priorities from ambient benzene to underground benzene plumes and occupational health concerns. This finding aided in Thrive’s process of narrowing down and reducing the number of CBA terms, given multiple competing priorities (e.g., jobs, gentrification, and staying power).

The biggest strength of this project was the relationship and capacity building of our THRIVEair team. We combined the expertise of Clean Up Circle members who have been organizing together for years advocating for remediation of the site, fenceline members with decades of lived experience in S/SW Philly, and Drexel scientists bringing scientific knowledge. One of the lead scientists on this project is also a long-time Thriver and member of the Clean Up Circle, which greatly helped to develop these relationships. By hiring Thrivers as community scientists through air monitoring and report-back co-design facilitation, we were also able to redistribute some funds into the community.

We successfully collaborated on domain and monitoring site selection through an iterative community mapping process, utilizing an “action-reflection-action” approach, to arrive at our final design and create dialogue around where Thrivers were most concerned about air pollution in their area, using a GIS-based web app to identify potential locations, updating locations between meetings based on feedback, and repeating this process.^20^

One of the limitations of this study is that we were unable to monitor benzene in real-time, which limited our ability to interpret short term VOC peaks. While several low-cost monitors can measure total VOC concentrations in real time, they lack the accuracy to detect individual VOC species at low concentrations.^21^ Notably, EPA regulatory monitoring in the US collects 24-hour samples using SUMMA canisters. While this approach offers a finer temporal resolution than our method, it cannot measure variation within a day.^22^

The results of this study pointed to a decrease in benzene concentrations over time as the former refinery was decommissioned. However, spatial variation in our measured benzene concentrations points to the continued need for air monitoring. Construction has already begun on the former refinery site, including a 25-year plan for a massive, multi-tenant logistics hub and life sciences campus. Thrivers are concerned about future increases in traffic-related air pollution, including benzene, as the HRP site continues to transition from a refinery to a warehousing/logistics center. Increases in air and sound pollution have been observed in environmental justice communities across the country due to the construction of warehouses.^23^^24^^25^

## Conclusion

THRIVEair Circle members designed an air monitoring campaign to track VOCs in neighborhoods adjacent to the former PES refinery and enhance data democratization by providing publicly available air pollution data in South/Southwest Philadelphia. This project is an example of how an environmental justice organization’s experience on the receiving end of government and corporate public participation processes can be used to improve data dissemination. We found that benzene and other VOC concentrations were generally low; however, spatial and temporal variation highlight the need for continued air monitoring. In particular, Thrivers have emphasized the need for real-time air monitoring results to inform day-to-day decision-making—while real-time monitoring is not feasible at scale for benzene and specific VOC species, total VOCs and PM_2.5_ can be monitored continuously.

Since the completion of THRIVEair in June 2024, several EPA-funded environmental justice grants have allowed other community organizations in Philadelphia to begin using low-cost sensors to monitor traffic-related air pollution (e.g., particulate matter) in real time. One of these projects will also continue to collaborate with a subset of THRIVEair Circle members to monitor benzene using the methodology developed in this project for VOC detection, in addition to real-time particulate matter monitoring.

## Data Availability

All data produced in the present study are available online at thriveairphilly.com or upon request to the authors

https://www.thriveairphilly.com

## Author Contributions

**Conceptualization:** ST, LF, KM, JN, SS, SA, CW, CF, DR, MC, CH, DH, JM, PW, CJ, JEC; **Data curation:** ST, LF, KM, JN, SA, GT, KW, LJ, NJ; **Formal analysis:** LF, KM; **Funding acquisition:** ST, JEC; **Investigation:** ST, LF, KM, JN, SS, SA, GT, KW, LJ, NJ; **Methodology:** ST, LF, KM; **Project administration:** ST, NJ, JEC; **Resources:** ST, LF, NJ, JEC; **Software:** LF, KM, NJ; **Supervision:** ST, LF; **Validation:** ST, KM, KW, LJ, NJ; **Visualization:** ST, LF, KM; **Writing – original draft:** ST, KM; **Writing – review & editing:** ST, LF, KM, PW, CJ, NJ, JM, JEC.

## Acknowledgements

The authors would like to thank the National Institute of Environmental Health Sciences (NIEHS grant # 5R21ES034494) Drexel Environmental Collaboratory, Drexel Peace Engineering/Build Up and the National Institute of General Medical Sciences of the National Institutes of Health (grant # P20GM103408)

## Footnotes

1 “The Chemical That Could Have Killed Millions in the Philly Refinery Fire,” *WHYY*, July 30, 2019, https://whyy.org/episodes/the-chemical-that-could-have-killed-millions-in-the-philly-refinery-fire/.

2 Susan Phillips and Dana Bate, “Faulty, Old Pipe Caused PES Refinery Explosion, Sending a Bus-Size Piece of Debris Flying across Schuylkill,” *WHYY*, October 16, 2019, https://whyy.org/articles/faulty-old-pipe-caused-pes-refinery-explosion-sending-a-bus-size-piece-of-debris-flying-across-schuylkill/.

3 “HRP Group Unveils The Bellwether District,” October 29, 2021, https://hilcoglobal.com/news/hrp-group-unveils-the-bellwether-district/.

4 Environmental Integrity Project, “Environmental Justice and Refinery Pollution: Benzene Monitoring Around Oil Refineries Showed More Communities at Risk in 2020,” April 28, 2021, https://environmentalintegrity.org/reports/environmental-justice-and-refinery-pollution/.

5 Poonam Kumari, Daya Soni, and Shankar G. Aggarwal, “Benzene: A Critical Review on Measurement Methodology, Certified Reference Material, Exposure Limits with Its Impact on Human Health and Mitigation Strategies,” *Environmental Analysis, Health and Toxicology* 39, no. 2 (June 2024): e2024012-2024010, https://doi.org/10.5620/eaht.2024012; Martyn T. Smith, “Advances in Understanding Benzene Health Effects and Susceptibility,” *Annual Review of Public Health* 31, no. 1 (March 1, 2010): 133–48, https://doi.org/10.1146/annurev.publhealth.012809.103646; Centers for Disease Control (CDC), “Benzene Chemical Fact Sheet,” 2024, https://www.cdc.gov/chemical-emergencies/chemical-fact-sheets/benzene.html.

6 Centers for Disease Control (CDC), “Benzene Chemical Fact Sheet.”

7 Amy Auchincloss, Kathleen Escoto, and Anneclaire De Roos, “Statement on the Health Effects of Refineries and Implications for the S Philadelphia Refinery” (Drexel University Dornsife School of Public Health, October 28, 2019).

8 Lars Barregard, Erik Holmberg, and Gerd Sallsten, “Leukaemia Incidence in People Living Close to an Oil Refinery,” *Environmental Research* 109, no. 8 (November 2009): 985–90, https://doi.org/10.1016/j.envres.2009.09.001; Cheng-Kuan Lin et al., “Lung Cancer Mortality of Residents Living near Petrochemical Industrial Complexes: A Meta-Analysis,” *Environmental Health* 16, no. 1 (December 2017): 101, https://doi.org/10.1186/s12940-017-0309-2; Tzu-Hsuen Yuan et al., “Increased Cancers among Residents Living in the Neighborhood of a Petrochemical Complex: A 12-Year Retrospective Cohort Study,” *International Journal of Hygiene and Environmental Health* 221, no. 2 (March 2018): 308–14, https://doi.org/10.1016/j.ijheh.2017.12.004.

9 Environmental Protection Agency, “The Benzene Fenceline Monitoring Dashboard,” Government, n.d., https://awsedap.epa.gov/public/extensions/Fenceline_Monitoring/Fenceline_Monitoring.html?sheet=background.

10 Environmental Protection Agency.

11 OAR US EPA, “Interactive Map of Air Quality Monitors,” Data and Tools, August 17, 2016, https://www.epa.gov/outdoor-air-quality-data/interactive-map-air-quality-monitors.

12 “Right to Thrive Campaign (2020 - Present),” Philly Thrive, 2020, https://www.phillythrive.org/campaign.

13 Meredith Minkler and Nina Wallerstein, *Community-Based Participatory Research for Health*, Third Edition (San Francisco: Jossey-Bass/Wiley, 2003).

14 Sarah L Fraser, “What Stories to Tell? A Trilogy of Methods Used for Knowledge Exchange in a Community-Based Participatory Research Project,” *Action Research* 16, no. 2 (June 2018): 207–22, https://doi.org/10.1177/1476750316680722.

15 Yoshira Ornelas Van Horne et al., “An Applied Environmental Justice Framework for Exposure Science,” *Journal of Exposure Science & Environmental Epidemiology* 33, no. 1 (2023): 1–11, https://doi.org/10.1038/s41370-022-00422-z.

16 Environmental Protection Agency, “The Benzene Fenceline Monitoring Dashboard.”

17 Dylan D. Miller et al., “Diffusive Uptake Rates for Passive Air Sampling: Application to Volatile Organic Compound Exposure during FIREX-AQ Campaign,” *Chemosphere* 287 (January 2022): 131808, https://doi.org/10.1016/j.chemosphere.2021.131808.

18 Gabrielle N. Dickinson et al., “Health Risk Implications of Volatile Organic Compounds in Wildfire Smoke During the 2019 FIREX-AQ Campaign and Beyond,” *GeoHealth* 6, no. 8 (August 2022), https://doi.org/10.1029/2021GH000546.

19 Evergreen Resources Group, LLC, *Summary of Area of Interest 11 (AOI 11)* (2020), https://phillyrefinerycleanup.info/act-2-documents/.

20 Minkler and Wallerstein, *Community-Based Participatory Research for Health*.

21 Milagros Ródenas García et al., “Review of Low-Cost Sensors for Indoor Air Quality: Features and Applications,” *Applied Spectroscopy Reviews* 57, no. 9–10 (November 26, 2022): 747–79, https://doi.org/10.1080/05704928.2022.2085734.

22 US EPA, “Interactive Map of Air Quality Monitors.”

23 Quan Yuan, “Location of Warehouses and Environmental Justice,” *Journal of Planning Education and Research* 41, no. 3 (September 2021): 282–93, https://doi.org/10.1177/0739456X18786392.

24 Jenni A. Shearston et al., “Opening a Large Delivery Service Warehouse in the South Bronx: Impacts on Traffic, Air Pollution, and Noise,” *International Journal of Environmental Research and Public Health* 17, no. 9 (May 5, 2020): 3208, https://doi.org/10.3390/ijerph17093208.

25 Priyanka N. deSouza, Sudheer Ballare, and Deb A. Niemeier, “The Environmental and Traffic Impacts of Warehouses in Southern California,” *Journal of Transport Geography* 104 (October 2022): 103440, https://doi.org/10.1016/j.jtrangeo.2022.103440.

