## Supplemental for "THRIVEair: A community-based air monitoring network design in a pollution-burdened Philadelphia neighborhood to advance environmental justice"

### Supplemental Figures

***Table of Contents***

**Figure S1.** Philadelphia Energy Solutions (PES) refinery benzene data…………………………..2

**Figure S2**. Philly Thrive 2022 action arc ..........................................................................3

**Figure S3.** Study domain selection…………………………………………………………….....4

**Figure S4.** THRIVEair source classes……………………………………..……………………......4

**Figure S5.** Information tags……………...…………………………….……..……………………......5

**Figure S6.** Benzene fact sheet……………......…………………………….……..……………………......5

**Figure S7.** Study design fact sheet……………......……………………..……...……………………......6

**Figure S8.** R Shiny App Screenshots……………......……………………..……...……………………......7

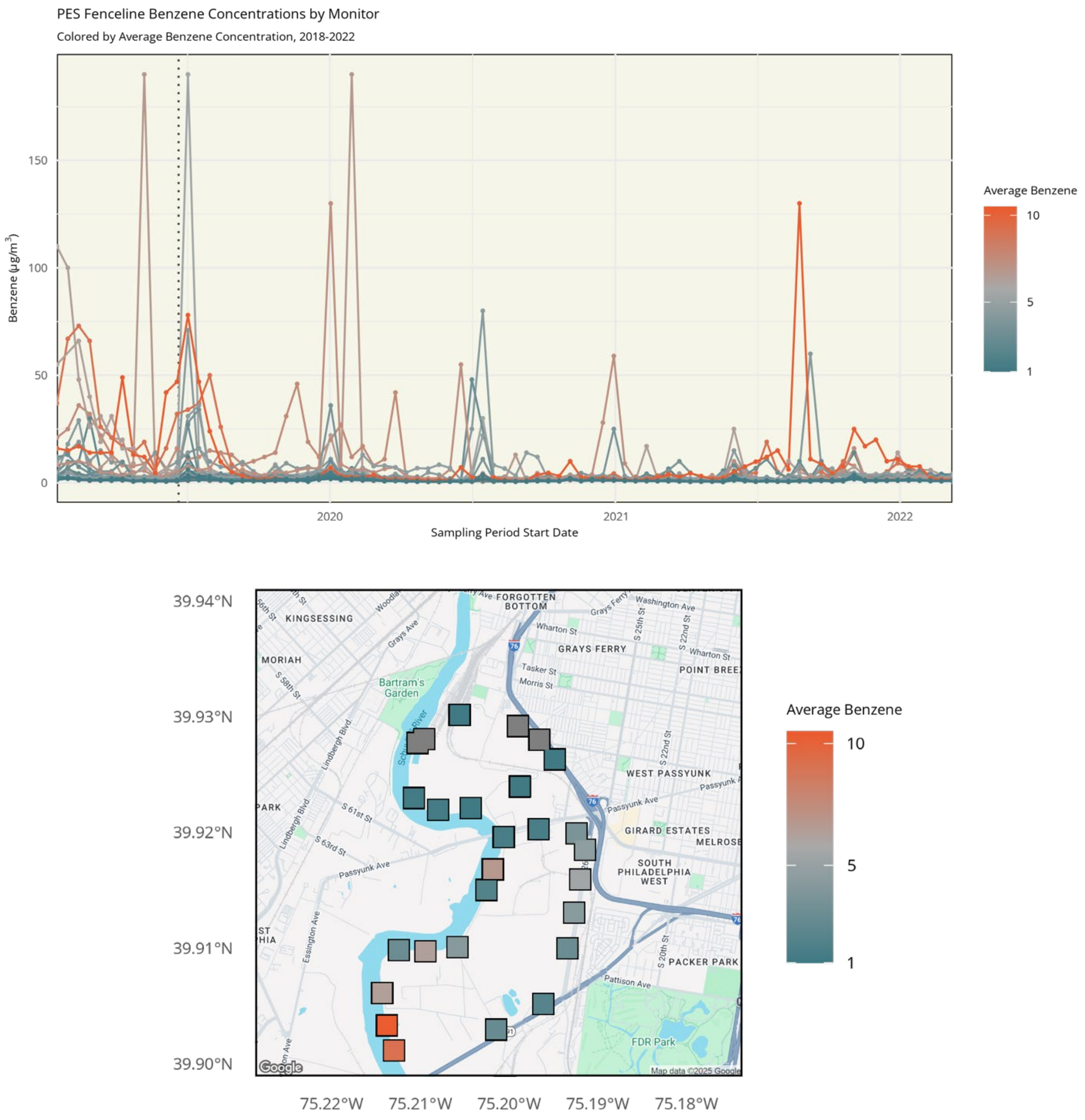


**Figure S1.** Philadelphia Energy Solutions (PES) refinery benzene data (2-week sampling sessions) reported to the US EPA from 2018-2022 by individual benzene monitor (top), and monitor site locations (bottom), colored by average benzene across the time period.

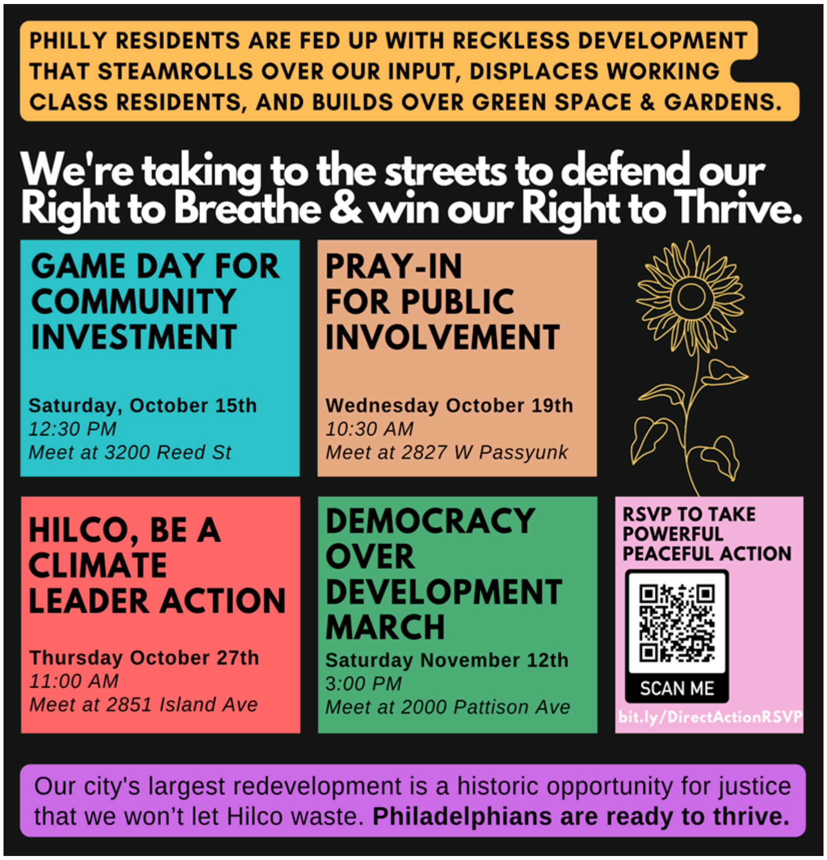


**Figure S2.** Philly Thrive's four-part non-violent direct action arc aimed to put pressure on HRP to negotiate a community benefits agreement through a public participation process. The third protest "Hilco be a Climate Leader" was in opposition to Philadelphia Air Management Services' announcement confirming the approval of HRP's permit to continue operations of the Schuylkill River tank farm.

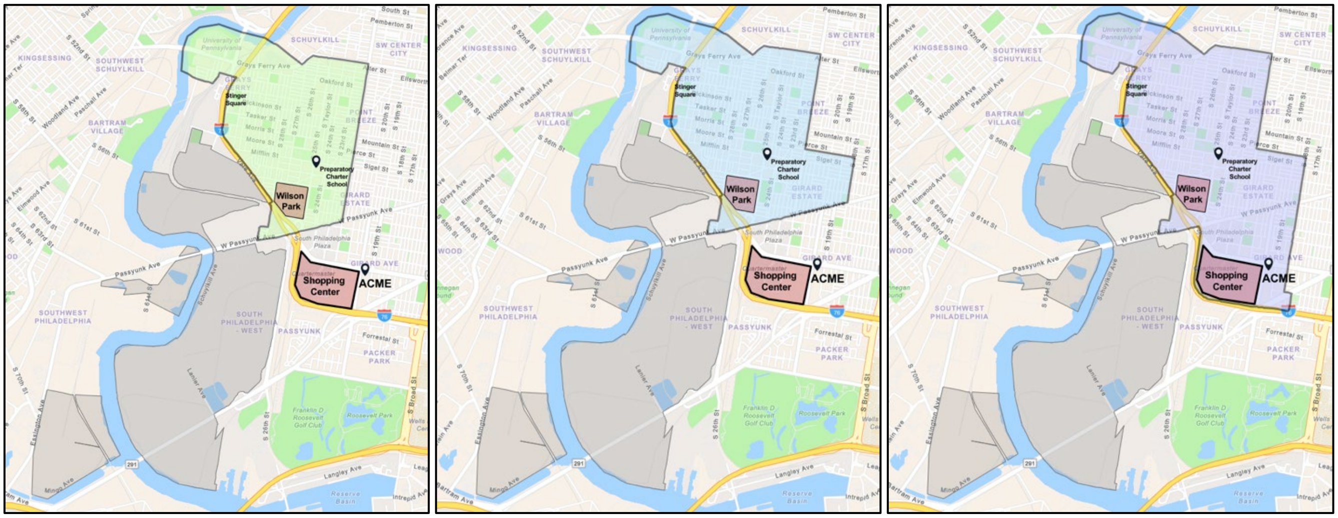


**Figure S3.** Study domain selection: 3 community domains presented to study design team.

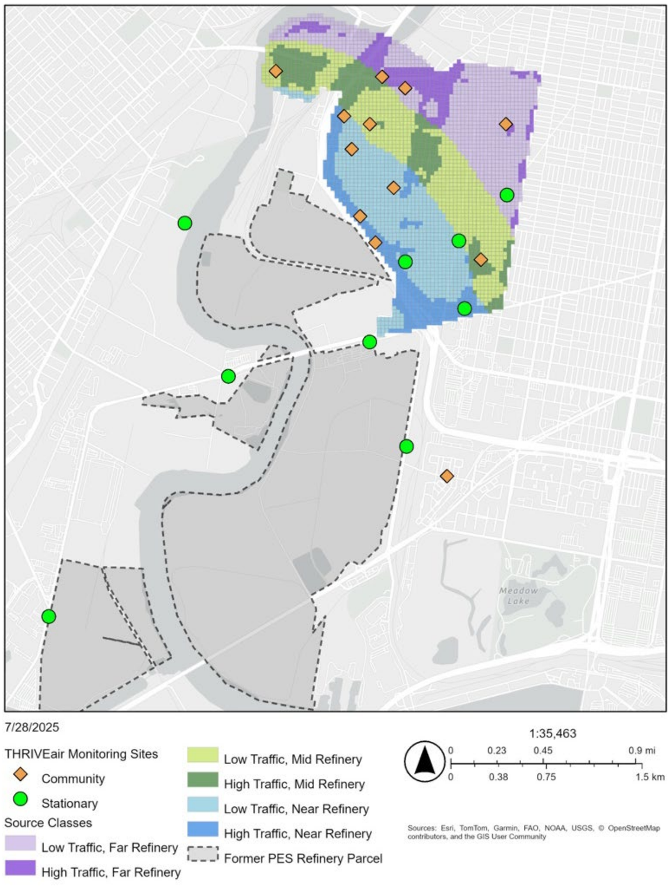


**Figure S4.** THRIVEair sources classes used for monitoring site selection in the community study domain.

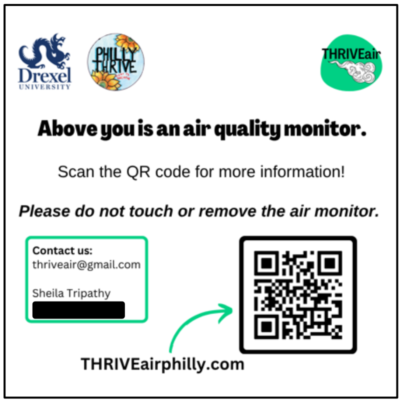


**Figure S5.** Information tags included underneath each air monitor. Contact phone number is redacted.

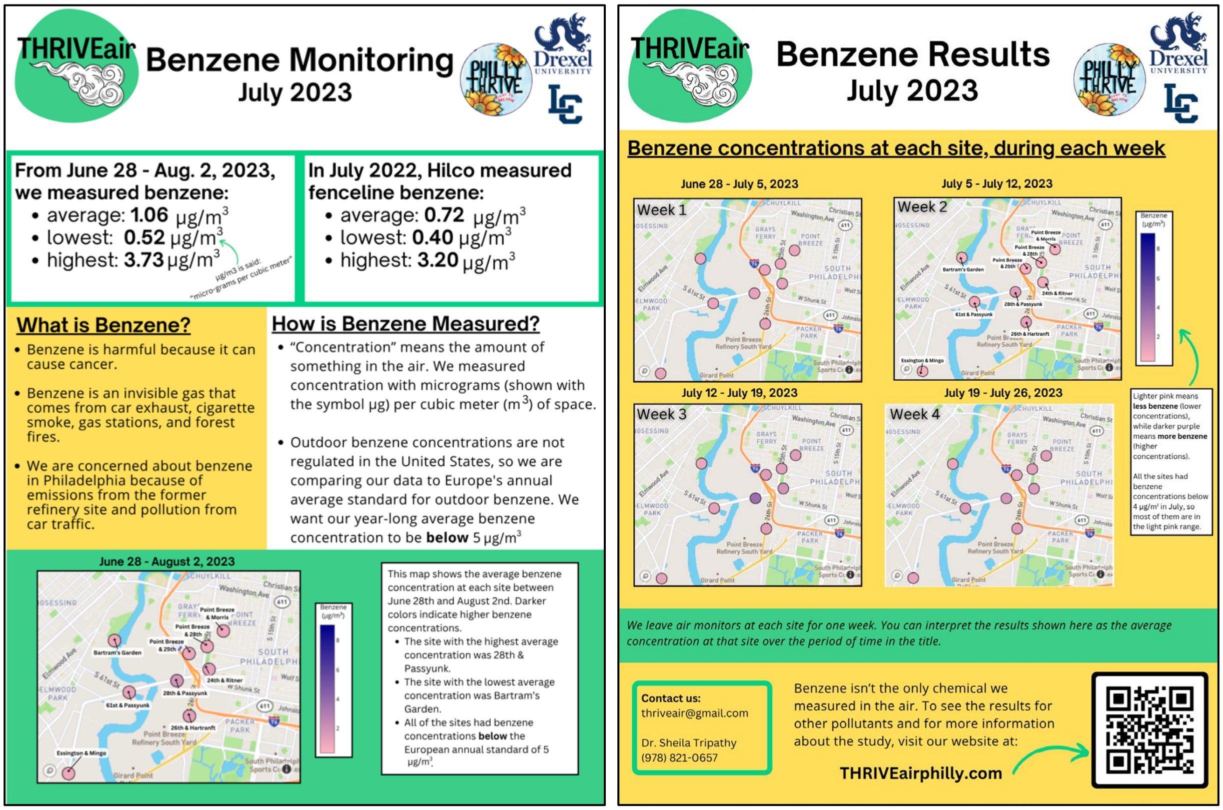


**Figure S6.** THRIVEair benzene fact sheet example.

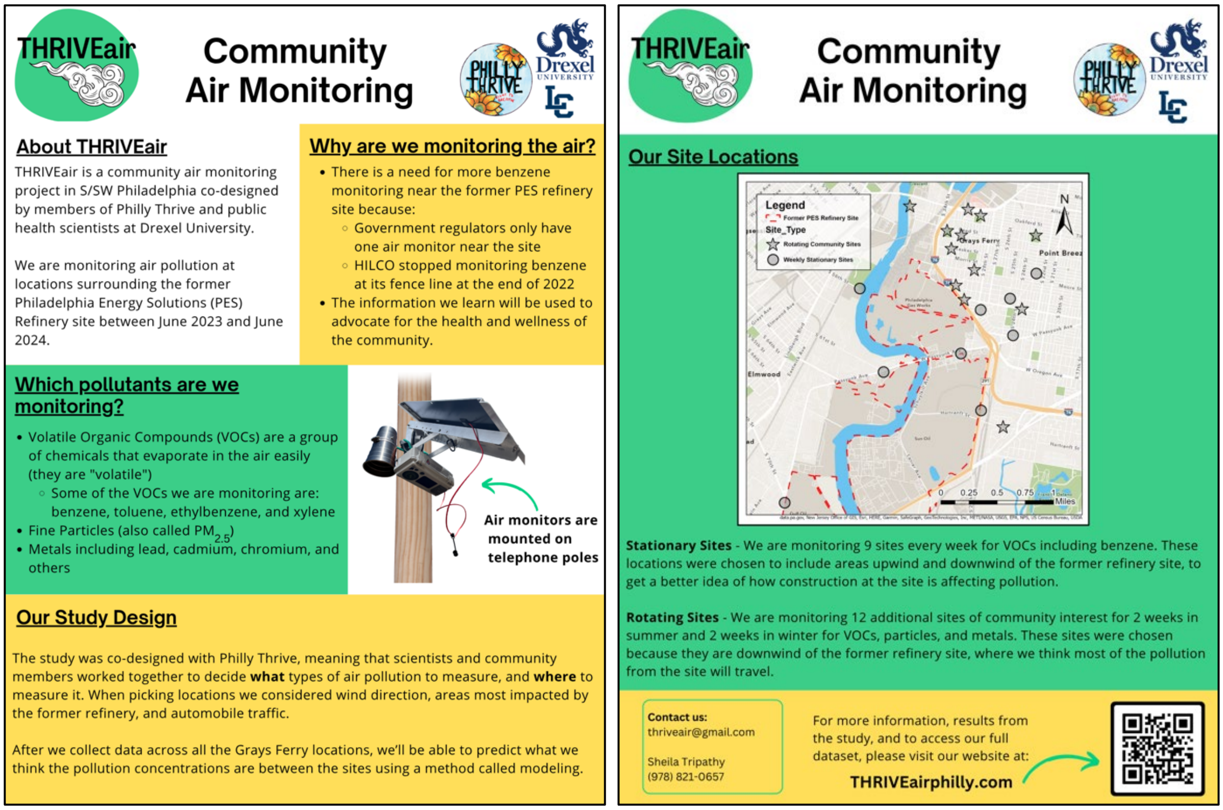


**Figure S7.** THRIVEair study design fact sheet.

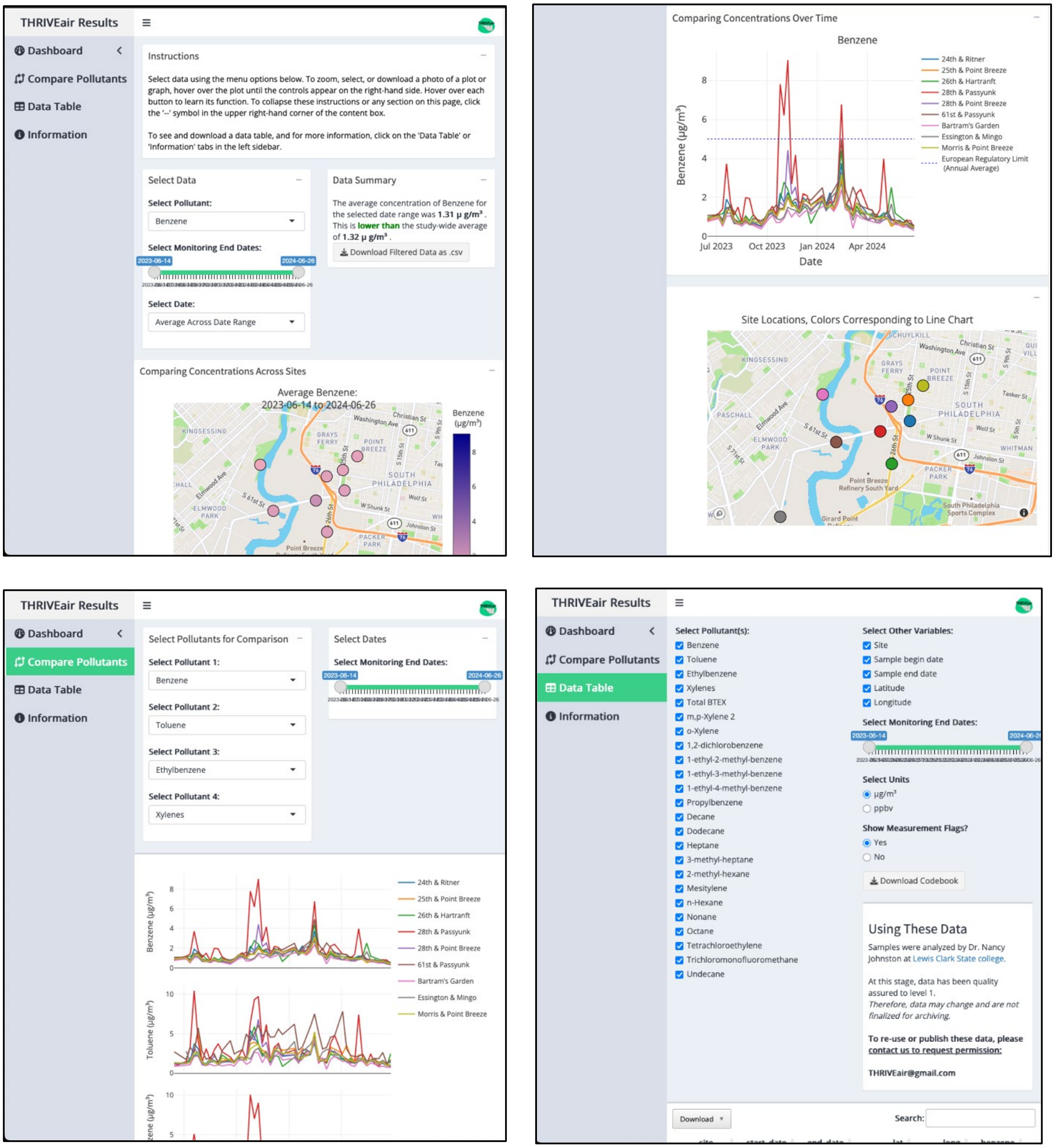


**Figure S8.** Screenshots from the thriveairphilly.com interactive R Shiny app.
